# An observational study examining nasopharyngeal resistome evolution among children under the selective pressure of antibiotic exposure and hospitalisation for pneumonia in Malawi: A study protocol

**DOI:** 10.64898/2026.11.11.26367839

**Authors:** Lucy O’Connor, Akuzike Kalizang’oma, Yamikani Chimalizeni, Conor Meehan, Grace Androga, Comfort Brown, Marc Henrion, Chrispin Chaguza, Neil French, Robert S. Heyderman, Brenda Kwambana-Adams

## Abstract

**Introduction:** Antimicrobial resistance (AMR) to first line antibiotics for pneumonia is a major global health problem, and AMR-attributable deaths are prevalent in sub-Saharan Africa. Improved data on how AMR develops and influences health outcomes in resource limited settings is crucial. The nasopharynx is a reservoir for pneumonia pathogens and their resistome, the genetic determinants of AMR. No study has explored whether human resistome composition in children with pneumonia is associated with health outcomes. This study will examine whether antibiotic treatment and hospital admission for pneumonia generate a persistent hospital-acquired nasopharyngeal resistome in children, and whether this is associated with re-hospitalisation.

**Methods:** This is an observational case-control study, that will recruit 350 children aged 12-36 months from four groups in Blantyre, Malawi: healthy children in the community (healthy community), children with pneumonia treated in the community (community pneumonia), children hospitalised with pneumonia (hospital pneumonia) and children re-hospitalised with pneumonia (re-hospitalised pneumonia). Nasopharyngeal swabs will be taken at recruitment and 3-month follow-up study visits for metagenomic sequencing to characterise the nasopharyngeal microbiome and resistome; urine samples will be taken at recruitment for an antibiotic activity bioassay. Socio-demographic data, co-morbidities, pneumonia severity, antibiotic exposure and health outcomes will be recorded.

**Analysis:** Cross-sectional comparisons between groups at recruitment will examine whether nasopharyngeal resistome (NPR) composition differs by pneumonia severity, antibiotic and healthcare exposure. Longitudinal comparisons within groups will assess the dynamics of NPR composition over time. The study is powered to detect a medium effect size (Cohen’s D = 0.505) change in the relative abundance of antibiotic resistance genes (ARGs) between all four groups. Primary outcomes are the difference in beta diversity (Bray-Curtis dissimilarity index), and relative abundance of ARG classes in the NPR. The primary objectives are to estimate the difference in diversity and relative abundance of the NPR between hospitalised pneumonia participants when compared to 1) healthy community and 2) community pneumonia participants, and to investigate whether participants re-hospitalised with pneumonia within a 3-month period have a persistent hospital-acquired NPR pattern.

**Ethics and dissemination:** The study has been approved by the Kamuzu University of Heath Sciences Research Ethics Committee (KUREC, formerly Malawi College of Medicine Research Ethics Committee; Ref: P.10/24-1200) and Liverpool School of Tropical Medicine Research Ethics Committee (LSTM REC; Ref: 24-076).

**Trial registration number:** ISRCTN64027792

**STRENGTHS AND LIMITATIONS OF THIS STUDY:** *Strengths:*

- Study design that uses metagenomic deep sequencing to enable direct, concurrent comparison of nasopharyngeal microbiome (NPM) and resistome (NPR) composition between community and hospitalised paediatric populations with pneumonia in a resource-limited setting.
- Data on reported and urinary bioassay-detected antibiotic use prior to, and throughout the study, enabling detailed examination of antibiotic selective pressure on the NPR.
- Investigation of the association between NPR composition and adverse clinical outcomes from pneumonia.

*Limitations:*

- Limitations to this study include few sampling time points (two for community pneumonia and three for hospitalised pneumonia), which may not fully capture the impact of antibiotic exposure; the potential for residual confounding following adjustment for known potential confounders (e.g. age, sex, co-morbidities, household exposure to smoke and breastfeeding) in the association between resistome composition and adverse clinical outcomes, for which the authors have detailed statistical analysis measures; and ability to only determine association between NPR composition and adverse clinical outcomes, rather than causal inference, due to the case-control study design.

**SUMMARY:** *What is already known on this topic:* The nasopharynx is a reservoir for pneumonia pathogens and their genetic determinants of antibiotic resistance in children. Studies of nasopharyngeal microbiome composition have shown association with paediatric pneumonia severity and mortality.

*What this study adds:* This study will describe nasopharyngeal resistome (NPR) evolution under selective pressure of antibiotics and hospitalisation for paediatric pneumonia, and explore whether NPR composition is associated with health outcomes in a resource-limited setting.

*How this study might affect research, practice or policy:* This exploratory study could inform larger scale observational studies to examine the association between antibiotic resistance and health outcomes, by defining high-risk nasopharyngeal resistome composition, helping to determine the required sample size and contributing open-source bioinformatic pipelines. The study could also inform research for public health interventions, including antibiotic stewardship.

## INTRODUCTION

Antimicrobial resistance (AMR) is a threat to global public health, predicted to cause 10 million deaths annually by 2050 (1). Although sub-Saharan Africa (sSA) is reported to be the region most affected by AMR-attributable death, these models are based on little empirical AMR data from the region (2). Therefore, to optimise public health interventions that reduce AMR-attributable morbidity and mortality, we need to improve our understanding of how antibiotic selective pressure and healthcare exposure influence AMR development in sSA, and how AMR impacts health outcomes.

Pneumonia is a leading cause of death in children under 5 years old worldwide (3) and a dominant syndrome in the global burden of AMR (2). The nasopharynx is a reservoir for pneumonia pathogens, including *Streptococcus pneumoniae* and *Staphylococcus aureus*, which are common causes of AMR-attributable deaths (2,4). Nasopharyngeal colonisation by these pneumonia pathogens is a necessary precursor to lower respiratory tract infection (LRTI) (4). Consequently, the nasopharyngeal microbiome (NPM) is a good proxy for the lower respiratory tract microbiome in paediatric pneumonia, with high concordance in microbiome composition between nasopharyngeal swabs (NPS) and deep respiratory tract samples in paediatric patients hospitalised with pneumonia (5).

The human respiratory microbiome is dominated by harmless colonising bacteria; pathogens represent just a fraction of the antibiotic resistance genes (ARGs; genetic code that is associated with antibiotic resistance mechanisms) found in these niches (6). However, bacteria have a remarkable ability to transfer genetic material within and between commensal and pathogenic genomes; ARGs in or on mobile genetic elements (MGEs; genetic code that moves between bacterial species) are the main drivers of AMR evolution, and pose the greatest risk to human health (7). For example, the nasopharynx is a conducive environment for horizontal gene transfer (HGT) of penicillin binding protein genes between *S. pneumoniae* and other streptococci (8,9).

Metagenomic deep-sequencing enables unbiased detection of the resistome, all the ARGs and MGEs in a microbial niche, analysis of which can unravel the complex evolutionary processes behind AMR, including decoding the origins of acquired, or extrinsic, ARGs and identifying intrinsic ARGs with inducible resistance. Examining nasopharyngeal resistome (NPR) evolution under selective pressure will enable the identification of populations at risk of antibiotic-resistant infections. This may inform response strategies through public health interventions, such as hospital admission policies and prescribing practices, including altered use of antibiotics to exploit synergistic combinations and enhance their bactericidal effects (10).

However, a major challenge of using metagenomics to study the NPR is the low bacterial biomass and high concentrations of human host DNA in nasopharyngeal samples; the larger human genome can completely obscure bacterial reads, making assembly challenging (10). Consequently, there are few studies of NPR evolution under antibiotic selective pressure, and existing studies have key limitations (see table 2). Importantly, whilst there are studies that show an association between respiratory microbiome composition and mortality in paediatric pneumonia (11), to date no study has examined whether human respiratory resistome composition is associated with health outcomes.

**Table 1.**
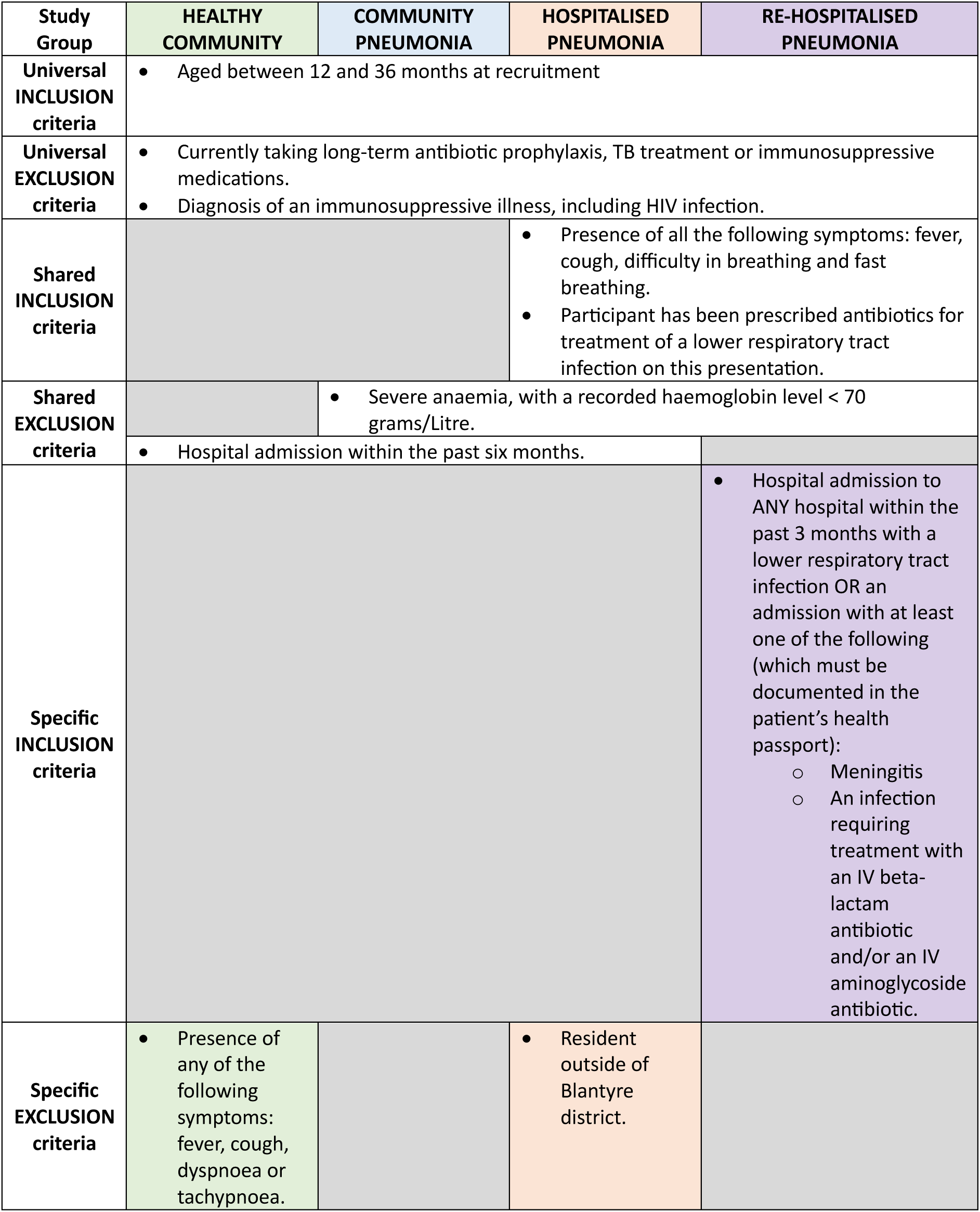
Inclusion and Exclusion Criteria for the study groups.

| **Study Group** | **HEALTHY COMMUNITY** | **COMMUNITY PNEUMONIA** | **HOSPITALISED PNEUMONIA** | **RE-HOSPITALISED PNEUMONIA** |
|---|---|---|---|---|
| **Universal INCLUSION criteria** | • Aged between 12 and 36 months at recruitment |  |  |  |
| **Universal EXCLUSION criteria** | • Currently taking long-term antibiotic prophylaxis, TB treatment or immunosuppressive medications.<br>• Diagnosis of an immunosuppressive illness, including HIV infection. |  |  |  |
| **Shared INCLUSION criteria** |  |  | • Presence of all the following symptoms: fever, cough, difficulty in breathing and fast breathing.<br>• Participant has been prescribed antibiotics for treatment of a lower respiratory tract infection on this presentation. |  |
| **Shared EXCLUSION criteria** |  | • Severe anaemia, with a recorded haemoglobin level < 70 grams/Litre. |  |  |
|  | • Hospital admission within the past six months. |  |  |  |
| **Specific INCLUSION criteria** |  |  |  | • Hospital admission to ANY hospital within the past 3 months with a lower respiratory tract infection OR an admission with at least one of the following (which must be documented in the patient's health passport):<br>○ Meningitis<br>○ An infection requiring treatment with an IV beta-lactam antibiotic and/or an IV aminoglycoside antibiotic. |
| **Specific EXCLUSION criteria** | • Presence of any of the following symptoms: fever, cough, dyspnoea or tachypnoea. |  | • Resident outside of Blantyre district. |  |

**Table 2.**
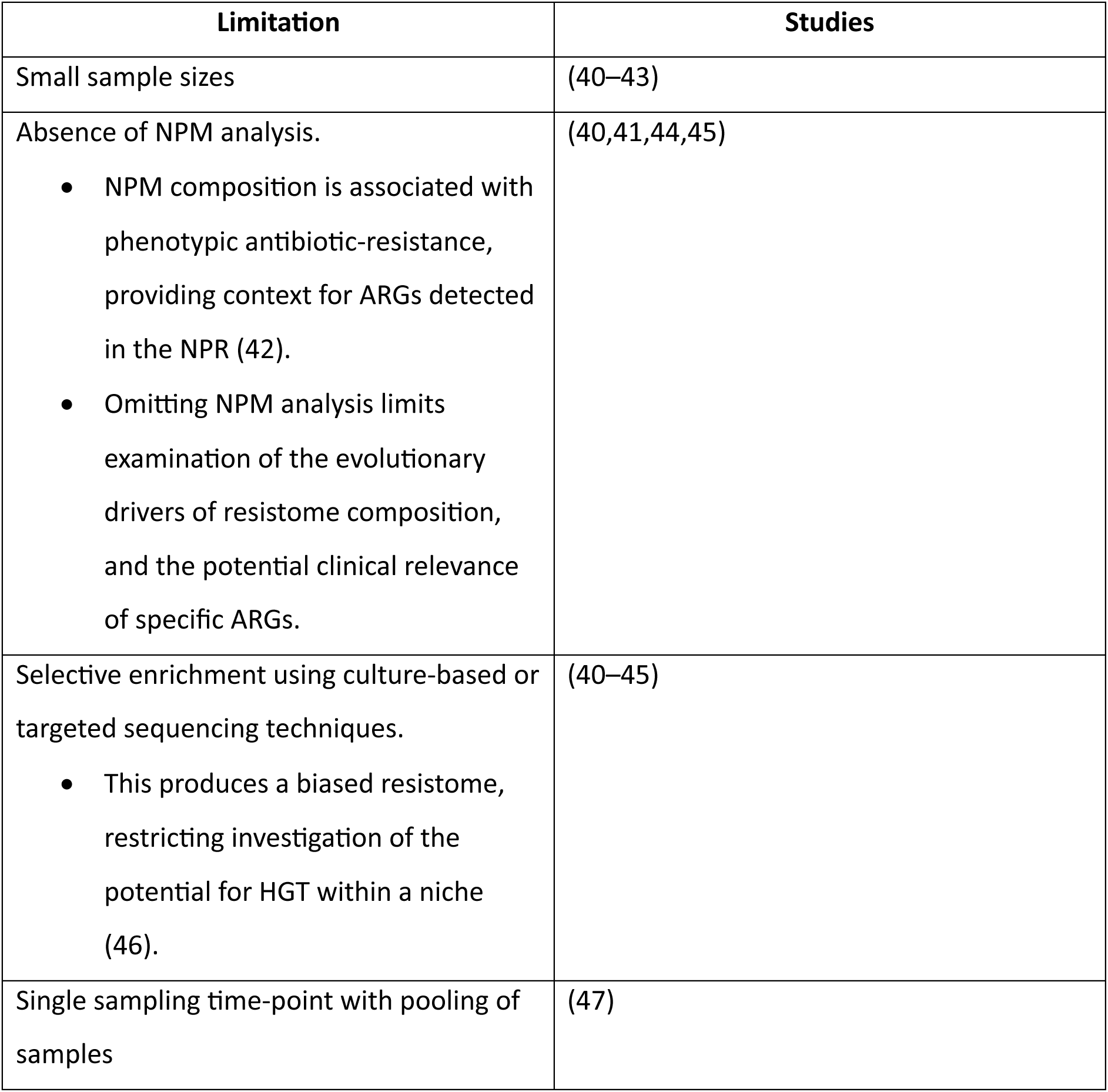
Limitations of existing studies of the NPR.

| **Limitation** | **Studies** |
|---|---|
| Small sample sizes | (40–43) |
| Absence of NPM analysis.<br>• NPM composition is associated with phenotypic antibiotic-resistance, providing context for ARGs detected in the NPR (42).<br>• Omitting NPM analysis limits examination of the evolutionary drivers of resistome composition, and the potential clinical relevance of specific ARGs. | (40,41,44,45) |
| Selective enrichment using culture-based or targeted sequencing techniques.<br>• This produces a biased resistome, restricting investigation of the potential for HGT within a niche (46). | (40–45) |
| Single sampling time-point with pooling of samples | (47) |

Malawi has limited healthcare resources and a high incidence of pneumonia in children aged 12-36 months (12). Current international guidance focuses on the identification of respiratory illness and provision of antibiotics (13); consequently, there is high antibiotic usage for children presenting with symptoms of a community-acquired respiratory tract infection (14,15). Our study aims to answer the following research question: does antibiotic treatment and hospital admission for pneumonia generate a persistent hospital-acquired NPR in Malawian children, and is this associated with adverse health outcomes in a resource-limited setting?

## METHODS AND ANALYSIS

### Study setting

The study will be conducted in Blantyre District, in southern Malawi, with an urban population of 1.3 million. Healthy children will be recruited from Ndirande Township, an informal settlement with a population of approximately 98,000 adjacent to Blantyre city (16). Children presenting with pneumonia will be recruited from Ndirande healthcare centre. Children hospitalised with pneumonia will be recruited from Queen Elizabeth Central Hospital (QECH), a government referral hospital providing free medical care to the peri-urban and rural residents of Southern Malawi (17).

### Study question

Does antibiotic treatment and hospital admission for pneumonia generate a persistent hospital-acquired NPR in Malawian children, and is this associated with adverse health outcomes in a resource-limited setting?

### Primary objectives

1. To measure how the diversity and relative abundance of the NPR of children hospitalised with pneumonia differs from the NPR of healthy children in the community.
2. To measure how the diversity and relative abundance of the NPR of children hospitalised with pneumonia differs from the NPR of children with pneumonia treated with antibiotics in the community.
3. To investigate whether participants re-hospitalised with pneumonia within a 3-month period have a persistent hospital-acquired NPR pattern.

### Secondary objective

4. To measure the impact of exposure to a hospital environment on the development of a ‘hospital-acquired’ NPR.

### Exploratory objective

5. To measure how the diversity and relative abundance of the NPR of children presenting with pneumonia differs between those with and without prior antibiotic use, based on urine bioassay.

### Study design

This is an observational case-control study with a cohort component to assess the evolution of the NPR under selective pressure of antibiotic use and hospital admission for pneumonia in children aged 12-36 months in Malawi. The study will describe the impact of antibiotic selective pressure and exposure to the hospital environment on the NPR of Malawian children with pneumonia. The study will also investigate the hypothesis that a hospital-acquired NPR is associated with adverse health outcomes following discharge, by examining whether NPR composition is associated with hospital readmission.

This study will recruit 350 children aged 12-36 months in four groups: healthy children in the community, children with pneumonia managed in the community, children hospitalised with pneumonia, and children re-hospitalised with pneumonia. For the healthy community group, 83 healthy children will be recruited from Ndirande Township using established recruitment protocols for pneumococcal carriage surveys (18); random number generation will be used to select the health surveillance assistant (HSA) catchment areas on a pre-defined map of Ndirande. The HSA will accompany the study field team to the selected geographical area over 2 days to identify households with age-appropriate candidates for recruitment. These healthy children will have a nasopharyngeal swab (NPS) sample collected at their recruitment study visit, to determine the baseline community resistome through metagenomic sequencing (see Figure 1). Healthy community recruitment will be conducted over 12 months to capture seasonal variation in NPM composition (19). This healthy community cohort will not have any follow-up study visits.

**Figure 1.**
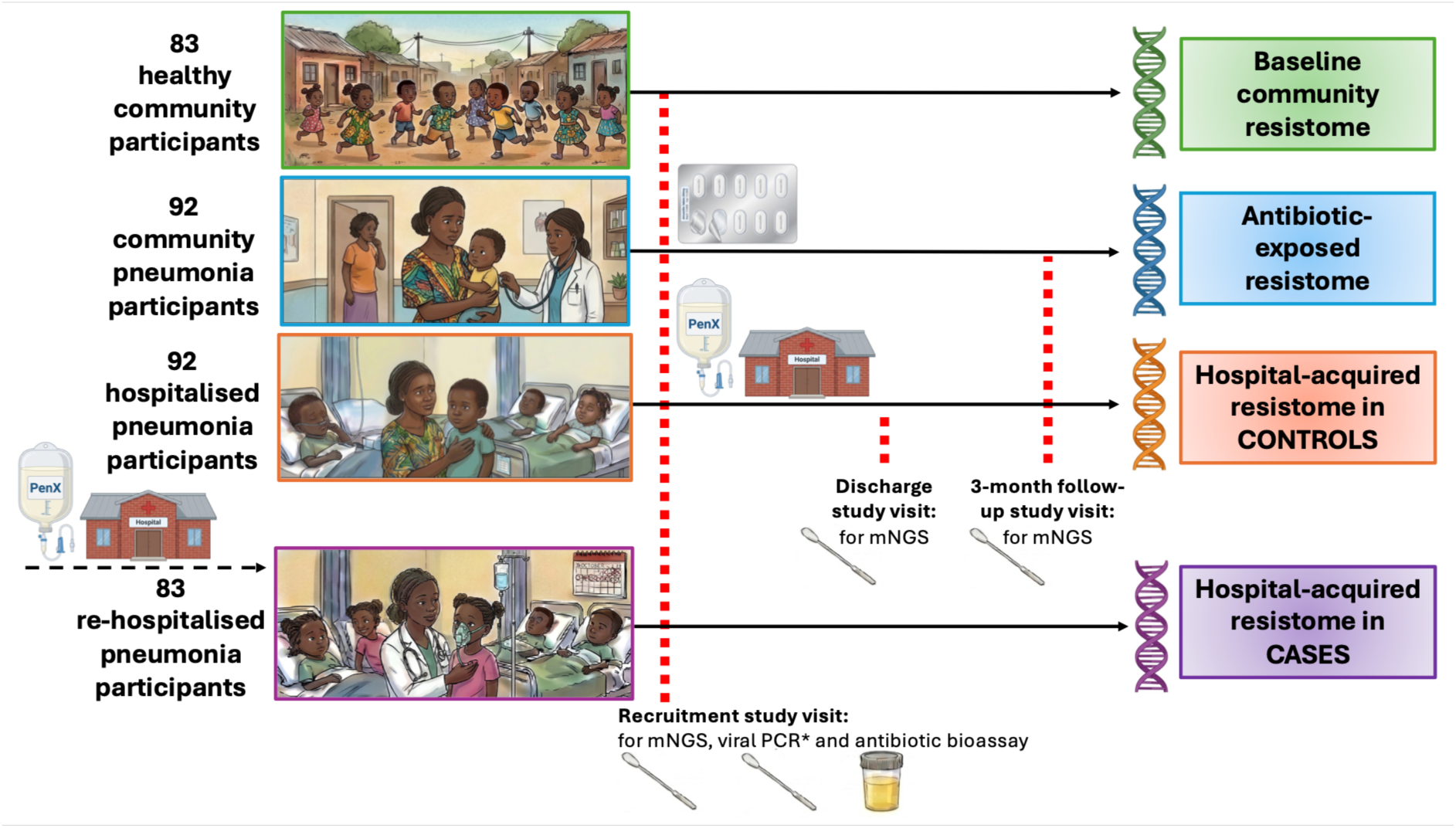
Study design and sampling frame. Red dashed lines show study visits. Small icons show antibiotic and hospital exposure. Hospitalised pneumonia participants (orange boxes) that are not re-hospitalised within 3 months of discharge form the control group for the case-control comparison. *Only participants with pneumonia will have a second NPS taken for viral PCR at recruitment. mNGS, shotgun metagenomic sequencing. Illustrations were generated using FigureLabs (48) and edited using Google’s Gemini 3 model (49).

92 children presenting with pneumonia to Ndirande healthcare centre, who are being prescribed antibiotic therapy in the community, will be recruited at the time of presentation to examine the impact of oral antibiotics on the NPR; recruited community pneumonia participants will have NPS samples collected at recruitment and a second study visit 3-months after recruitment, to determine the antibiotic-exposed resistome (see Figure 1). Recruitment of healthy community and community pneumonia participants was conducted from August 2025 to July 2026.

92 children hospitalised with pneumonia at QECH will be recruited within 72 hours of admission to examine the impact of broad-spectrum intravenous antibiotics and exposure to a hospital environment on the NPR; these hospital pneumonia participants will have NPS samples collected at recruitment, hospital discharge and 3-month study visits to assess for a hospital-acquired resistome (see Figure 1).

The case-control component of the study will examine whether a hospital-acquired NPR is associated with adverse health outcomes. The study will focus on hospital re-admission as an adverse outcome related to AMR in pneumonia, because very severe pneumonia requiring hospitalisation is more commonly bacterial in aetiology (20); therefore, re-hospitalisation is more likely to reflect the impact of AMR in bacterial pneumonia. The following adverse health outcomes of AMR in bacterial pneumonia will also be recorded at 3-month study visits: recurrent episodes of pneumonia, additional courses of antibiotics for pneumonia and mortality outcomes.

Children hospitalised at QECH with pneumonia, who are not re-hospitalised within 3 months of recruitment will form the control group (see Figure 1); these participants will already have had a NPS collected at their 3-month study visit to assess for a hospital-acquired resistome.

83 children admitted to QECH with pneumonia within 3 months of a previous hospital admission for pneumonia will form the case group; these re-hospitalised pneumonia participants will be recruited within 72 hours of admission and have NPS samples collected at the recruitment study visit to assess for a hospital-acquired resistome. Health passport screening and carer-reporting will be used to establish prior hospitalisation. These re-hospitalised pneumonia participants will not have any follow-up study visits. Throughout this protocol, controls will be referred to as hospitalised pneumonia participants, and cases will be referred to as re-hospitalised pneumonia participants. Recruitment of hospitalised pneumonia and re-hospitalised pneumonia participants commenced in February 2026 and will be conducted over 12 months, or until the target sample size is achieved, if this is earlier.

A urine sample will be collected from all study participants at recruitment for a urine antibiotic activity bioassay to determine antibiotic consumption prior to enrolment (21).

### Inclusion and exclusion criteria

Inclusion and exclusion criteria are detailed in Table 1. Briefly, all study participants must be aged between 12 and 36 months at recruitment, cannot be on long-term antibiotic prophylaxis, nor treatment for tuberculosis or immunosuppressive medications, and must not have a known diagnosis of an immunosuppressive illness, including HIV infection; these factors may introduce potential confounding through altered NPM and NPR composition, or suppressed immune response to pneumonia. HIV is independently associated with mortality post-discharge in paediatric pneumonia (22).

In keeping with the standard of care in Malawi, pneumonia will be defined clinically using World Health Organization (WHO) criteria: reported or recorded fever, cough, dyspnoea and tachypnoea (13). Healthy community participants will be excluded if they have any symptoms of pneumonia.

To meet the inclusion criteria, participants with pneumonia (community pneumonia, hospitalised pneumonia and re-hospitalised pneumonia) must have all the symptoms of pneumonia, and be prescribed antibiotics for pneumonia. Parental reporting of fever based on tactile assessment has a good positive predictive value (23,24).

All participants except re-hospitalised participants will be excluded if they have had a hospital admission in the preceding 6 months. Conversely, hospital admission within the preceding 3 months with pneumonia, meningitis or an infection requiring intravenous beta-lactam and/or aminoglycoside antibiotic is an inclusion criterion for re-hospitalised pneumonia participants; this ensures that the antibiotic exposure in the preceding admission is always a beta-lactam and/or aminoglycoside antibiotic, based on the local and WHO guidance for these clinical syndromes. Participants with pneumonia will be excluded if they have severe anaemia (<70g/L), identified on tests conducted as part of routine clinical care; severe anaemia is a leading cause of recurrent hospitalisation in African children (25). Finally, hospital pneumonia participants must be resident within Blantyre, to facilitate attendance of the 3-month study visit.

### Sample collection

Healthy study participants will have one NPS taken at recruitment for metagenomic sequencing. Two NPS will be taken at the recruitment study visit from all study participants with pneumonia; one NPS will undergo metagenomic sequencing, the other NPS will be tested for respiratory viruses using polymerase chain reaction (PCR) to identify the pathogens most commonly associated with severe acute respiratory infections requiring hospitalisation in the paediatric population in Malawi: Respiratory Syncytial Virus (RSV), Influenza A and Influenza B (26). Viral infections are a potential confounder in the association between NPR composition and re-hospitalisation with pneumonia. One NPS will be taken from study participants at subsequent study visits: at discharge from hospital and the 3-month follow-up visit. All collected NPS will be transported at 2-8°C from the field to the Malawi Liverpool Wellcome Research Programme (MLW) laboratory in Blantyre, where they will be processed and stored at −80°Celsius, following an established protocol (18), prior to DNA extraction and metagenomic sequencing. Sweep cultures of NPS on Sheep’s Blood Agar (SBA) and Chocolate agar, and *S.pneumoniae* isolates grown on SBA with gentamicin, will be stored at −80°Celsius.

A urine sample will be collected from all study participants at recruitment using a paediatric urine collection bag, transported at 2-8°Celsius from the field to the MLW laboratory and stored at −80°Celsius for a urine antibiotic activity bioassay to determine antibiotic use by study participants prior to recruitment (21). Reported antibiotic consumption data in the 7 days preceding recruitment will also be recorded, based on parent/carer reporting and health passport records; antibiotic consumption data will also be recorded at discharge and 3-month study visits to examine the impact of antibiotic selective pressure on the NPR.

### Urine antibiotic bioassay

The bioassay will detect antibiotic activity in urine samples collected from study participants at recruitment, by the presence of inhibited growth on Mueller-Hinton agar around a filter paper disc containing the urine sample of any of the following organisms: Geobacillus stearothermophilus (ATCC 7953), Streptococcus pyogenes (ATCC 19615) or Escherichia coli (ATCC 25922) on Mueller-Hinton agar (21).

### Metagenomic sequencing

Shotgun metagenomic sequencing will be performed on DNA extracted from liquid Amies media from NPS. All reads aligning to the human genome will be removed in the bioinformatic pipeline, which will be developed by the authors. Raw metagenomic reads will undergo quality checks and trimming, prior to assembly of metagenome-assembled-genomes, further quality checks, taxonomic profiling using the National Centre for Biotechnology Information database and ARG detection using the Comprehensive Antibiotic Resistance Database.

Negative controls will be included in sequencing to permit monitoring of contaminants throughout sample collection and processing: (i) sampling blank controls, to detect contaminant DNA introduced during sample collection, (ii) DNA extraction blank controls, to monitor for contaminant DNA content in extraction kits and sample processing and (iii) no-template amplification controls, to monitor contaminant DNA in reagents and the laboratory environment during library preparation and sequencing (Eisenhofer et al., 2019).

### Quality control for metagenomic sequencing data

The following pre-defined analysis thresholds will be applied to bacterial reads in the metagenomic sequence data to ensure high-confidence alignments to taxa present in the NCBI database and filter out extremely rare taxa, to remove possible contaminants, reduce extreme sparsity in microbiome data and technical variability from sample processing: >90% identity matches of reads and assembled contigs to reference sequences, alignment lengths ≥ 50bp to reduce false positive alignments, relative abundance ≥ 1% of reads per sample and rare taxa present in < *k* samples, where *k* is determined by 0.01% of the minimal library size. This improves reproducibility and comparability of microbiome results, whilst preserving beta diversity (27). A true-positive result will be valid only when the sequence reads exceed 10 times the corresponding reads in the negative control (28).

Bacterial reads will be normalised using Reads Per Kilobase per Million (RPKM) mapped reads, permitting quantitative comparison of the relative abundance of bacterial and ARG reads between samples, by dividing both transcript length and total number of reads (29).

### Expected outcomes

Primary outcomes are the composition of the NPR, as measured by the diversity and relative abundance of ARG classes detected by metagenomic sequencing of NPS samples collected from: hospitalised pneumonia participants at recruitment, discharge and 3-month follow up; community pneumonia participants at recruitment and 3-month follow up; healthy community participants at recruitment; re-hospitalised pneumonia participants at recruitment.

The primary endpoints for this study are:

1. The difference in beta diversity of ARG classes in the NPR, as measured by the Bray Curtis dissimilarity index, and difference in relative abundance of ARG classes in the NPR between healthy community and hospital readmission groups at enrolment, and community pneumonia and hospital pneumonia groups at the 3-month study end-point.
2. The difference in beta diversity of ARG classes in the NPR, as measured by the Bray Curtis dissimilarity index, and difference in relative abundance of ARG classes in the NPR between recruitment, discharge and 3-month study time-points in the hospital pneumonia group.
3. The difference in beta diversity of ARG classes in the NPR, as measured by the Bray Curtis dissimilarity index, difference in relative abundance of ARG classes in the NPR between recruitment and the 3-month study end-point in the community pneumonia group.

The first primary study endpoint will compare samples collected at the 3-month study visit for hospital pneumonia and community pneumonia participants, as these time-points will represent the new baseline resistome post-exposure, based on studies of NPM recovery following antibiotics for infective exacerbations of asthma in children (30). Beta diversity will be measured using the Bray-Curtis dissimilarity index; alternative indices of beta diversity may be explored.

This study will investigate the hypothesis that a hospital-acquired NPR is associated with adverse health outcomes following discharge by comparing NPR composition between participants readmitted to hospital with pneumonia within a 3-month period, and participants that remain in the community for 3-months after hospitalisation with pneumonia; the limitation of this comparison is that it cannot rule out reverse causation. Secondary outcomes include additional measures of NPR and NPM composition, based on the relative abundance of individual ARGs and bacterial species respectively, detected by metagenomic sequencing of NPS samples collected from study groups at study visit timepoints, as described for the primary outcomes. Secondary outcomes also include detection of viral pathogens RSV, Influenza A and B in recruitment NPS from participants with pneumonia; antibiotic use pre-enrolment for all study participants, measured by urinary antibiotic activity bioassay, or (if not available) reported antibiotic use; pneumonia severity at hospital admission; reported duration of hospital admission and shared bed-spaces for children hospitalised with pneumonia; reported hospitalisation, healthcare clinic attendance, antibiotic prescriptions and mortality at 3-month follow up of community pneumonia and hospital pneumonia participants.

Secondary endpoints are detailed in table 3. In brief these include: the difference in alpha diversity and relative abundance of ARGs in the NPR for each primary endpoint measured above; comparisons dependent on differences in primary endpoints for diversity and relative abundance of the NPR between discharge and 3-month study visits in the hospital pneumonia group, and recruitment and 3-month study visits in the community pneumonia group; the difference in diversity and relative abundance of bacterial taxa in the NPM for each primary endpoint listed above; the difference in the incidence of adverse health outcomes between community pneumonia and hospital pneumonia participants at 3-months.

**Table 3.** Study secondary endpoints. Antibiotic resistance genes, ARGs; nasopharyngeal resistome, NPR; nasopharyngeal microbiome, NPM.

| **Secondary Endpoints: NPR Analysis (Secondary outcomes)** |  |  |
|---|---|---|
| 1 | The difference in alpha diversity and relative abundance of individual ARGs in the NPR between healthy community and hospital readmission cohorts at enrolment, and community pneumonia and hospital pneumonia cohorts at the 3-month study end-point. |  |
| 2 | The difference in alpha diversity and relative abundance of individual ARGs in the NPR between recruitment, discharge and 3-month study time-points in the hospital pneumonia cohort. |  |
| 3 | The difference in in alpha diversity and relative abundance of individual ARGs in the NPR between recruitment and the 3-month study end-point in the community pneumonia cohort. |  |
| **Secondary Endpoints: NPR Analysis (Dependent on primary endpoint analyses)** |  |  |
| 4 | *(Dependent on primary study endpoint 2) If there is a change in NPR composition between discharge and the 3-month study end-point in the hospital pneumonia cohort:* The difference in the following measures of NPR composition between the hospital pneumonia cohort at discharge, the healthy community and hospital readmission cohorts at enrolment, and the community pneumonia cohort at the 3-month study end-point: | a) ARG richness and evenness (alpha diversity) of the NPR<br>b) Overall community composition (beta diversity) of the NPR<br>c) Relative abundance of ARG classes in the NPR<br>d) Relative abundance of individual ARGs in the NPR |
| 5 | *(Dependent on primary study endpoint 3) If there is a change in NPR composition between enrolment and the 3-month study end-point in the community pneumonia cohort:* The difference in the following measures of NPR composition between the community pneumonia cohort at enrolment, the healthy community and hospital readmission cohorts at enrolment, and the hospital pneumonia cohort at the 3-month study end-point: |  |
| 6 | *(Dependent on primary study endpoints 2 and 3) If there is a change in NPR composition between discharge and the 3-month study end-point in the hospital pneumonia cohort, and a change in the NPR composition between enrolment and the 3-month study end-point in the community pneumonia cohort:* The difference in the following measures of NPR composition between the healthy community, community pneumonia and hospital readmission cohorts at enrolment, and the hospital pneumonia cohort at discharge: |  |
| **Secondary Endpoints: NPM Analysis** |  |  |
| 7 | The difference in the following measures of NPM composition between the healthy community and hospital readmission cohorts at enrolment, and community pneumonia and hospital pneumonia cohorts at the 3-month study end-point: | a) Species richness and evenness (alpha diversity) of the NPM<br>b) Overall taxonomic composition (beta diversity) of the NPM<br>c) Relative abundance of bacterial taxa in the NPM |
| 8 | The difference in the following measures of NPM composition between recruitment, discharge and 3-month study time-points in the hospital pneumonia cohort: |  |
| 9 | The difference in the following measures of NPM composition between recruitment and the 3-month study end-point in the community pneumonia cohort: |  |
| **Secondary Endpoints: Health Outcomes** |  |  |
| 10 | The difference in the incidence of adverse health outcomes between the community pneumonia cohort and hospital pneumonia cohort at the 3-month study end-point: a) Repeat healthcare clinic attendances, b) repeat antibiotic prescriptions, c) hospitalisation and d) mortality. |  |

Exploratory analyses will investigate the association between NPR composition at discharge for hospitalised participants and recurrent hospitalisation with pneumonia; examine the compositional dissimilarity of the NPR between healthy community and hospital readmission cohorts at enrolment, and community pneumonia and hospital pneumonia cohorts at the 3-month study end-point based on hierarchical clustering of Bray-Curtis dissimilarity indices; explore the association between NPR composition based on hierarchical clustering of Bray-Curtis dissimilarity indices and recurrent hospitalisation with pneumonia; investigate the association between pneumonia severity and NPR composition at recruitment for hospitalised participants; estimate how the duration of hospitalisation and shared bed-spaces impact NPR composition at discharge in hospitalised pneumonia participants; estimate the impact of antibiotic use prior to recruitment, as detected by a urine bioassay, on NPR composition; and estimate differences in relative abundance of MGEs between recruitment and discharge time points in hospitalised pneumonia participants.

### Data collection

Demographic and clinical data will be collected on password-encrypted tablets using the electronic data capture system ODK (31). The following data on host and disease characteristics will be recorded for all study participants to assess for potential confounding in the association between NPR composition and re-hospitalisation with pneumonia: age, sex, co-morbidities, household exposure to smoke from tobacco or biomass fuels and exclusive breastfeeding for the first 3 months of life, as these variables are associated with pneumonia requiring hospitalisation in high-income settings (5); mid-upper arm circumference as an indicator of malnutrition, which is associated with pneumonia mortality (32); pneumococcal conjugate and *Haemophilus influenzae B* vaccination status, which are both associated with reduced incidence of bacterial pneumonia in this setting (14); and malaria rapid diagnostic test result and haemoglobin level, as severe anaemia is the leading cause of recurrent hospitalisation in children in Malawi (25).

There is a correlation between NPM composition, susceptibility to pneumonia and pneumonia severity in the paediatric population (33,34); consequently, determinants of NPM composition could be confounders in the association between NPR composition and hospital re-admission. To assess for this, we will record factors that influence early NPM development, including mode of childbirth (vaginal versus caesarean section) (35).

WHO markers of severity in pneumonia (13) at the time of admission will be recorded for hospitalised pneumonia participants, to describe the association between pneumonia severity, NPM and NPR composition; if heterogeneity exists, investigations will be performed for interaction and confounding in the association between NPR composition and recurrent hospitalisation with pneumonia. Pneumonia severity markers include central cyanosis, oxygen saturations less than 90%, inability to feed, vomiting, convulsions, lethargy and impaired consciousness. These severity markers are used in international and local Malawian guidelines to determine which paediatric patients with pneumonia should be referred to hospital. However, a recent study in northern Malawi showed that a minority (30.4%) of children hospitalised with pneumonia met these severity criteria, most admission decisions were based on hospital practices and health-seeking behaviour (14). To assess for the possible influence of healthcare-seeking behaviour on hospital re-admission, data will be collected on the highest educational level attained by the carer/parent and time taken to travel to the healthcare facility; these are major determinants of healthcare-seeking behaviour in Malawi (36).

Blood culture results and chest radiographs taken as part of routine clinical care for hospitalised study participants will be recorded. Whilst previous studies have shown a bacteraemia rate of less than 3% in hospitalised paediatric pneumonia in Malawi and sub-Saharan Africa (14,20), the antibiotic resistance profile of a pathogen isolated by blood culture from a study participant with pneumonia can be compared with the NPM and NPR from the same study participant. This will enable more detailed examination of the association between NPR composition, pneumonia aetiology and susceptibility to first line antibiotics for pneumonia in Malawi.

Chest radiographs are poorly predictive of aetiology in paediatric pneumonia and do not improve clinical outcomes (37). Consequently, WHO and local Malawian diagnostic guidelines only recommend chest radiographs for paediatric patients with severe pneumonia, to identify potential pneumonia complications including pleural effusions, empyema, pneumothoraces and pneumatoceles. Chest radiograph images taken during the admission in which hospitalised pneumonia and re-hospitalised pneumonia participants are recruited to the study, will be recorded to identify potential confounding in the association between NPR composition and hospital re-admission with pneumonia, by determining cases where re-hospitalisation could be attributed to complications of pneumonia rather than AMR.

To assess for acquisition of ARGs from the hospital environment, the following data will be recorded for hospitalised pneumonia participants: duration of hospitalization and incidence of shared bed-spaces, including number of bed occupants and number of days in a shared bed-space.

Community pneumonia and hospitalised pneumonia participants will have a 3-month study visit to their home, at which health outcomes including mortality, healthcare interactions (including healthcare centres and hospital attendance/admission) and antibiotic exposure will be recorded.

### Data management and anonymisation procedures

Each participant will be assigned a unique participant identification number (PID) at recruitment. NPS and urine samples will be labelled with a barcode linked to this PID; all study data, including laboratory and metagenomic sequencing results, will be pseudonymised using this PID. Pseudonymised data captured electronically on ODK will be transferred daily to a password-encrypted database using a secure connection at MLW, and backed up daily to local and off-site facilities, in compliance with Malawian data transfer legislation.

A logbook containing identifiable participant information will be kept separately in a secure location, accessed only by authorised study team members, to facilitate follow-up study visits. Study documentation, including consent forms, will be archived in-country for 10 years, as per the study sponsor’s policy.

### Sample size calculation

There is no established method for sample size calculation in resistome studies; as such the approach used here is for microbiome studies (38). Differences in gut plasmidome composition have been detected with 41 participants per group (39), but NPR has a lower biomass than gut resistome, therefore the resistome is likely to be less diverse, with a lower relative abundance of ARGs. NPR analysis 3.5 years post-antibiotic exposure in a cluster-randomised trial of mass-azithromycin administration showed a trend toward decreased NPR diversity (*p* = 0.07) in azithromycin clusters when compared to placebo clusters, with just 25 samples per group (unpublished data).

Using the measured difference in relative abundance of a specific macrolide ARG (ermX) 3.5 years after mass-azithromycin administration (unpublished data) as a medium effect size that may be clinically relevant (Cohen’s D = 0.505), accepting 90% power and 5% type I error (95% confidence interval), and applying a sample size calculation for a standard t test, as recommended for microbiome studies (38): n=2(Z_1-α/2_+ Z_1-β_)^2^/Δ^2^, the calculated sample size is 83 participants per group.

In total, the study aims to recruit 350 study participants: 83 healthy community participants, 92 community pneumonia participants, 92 hospital pneumonia participants and 83 re-hospitalised pneumonia participants. Study groups with a sample size of 92 participants permit a 10% loss to follow-up.

### Statistical analyses

Participants will be frequency-matched for sex across the healthy community, community pneumonia, hospitalised pneumonia and re-hospitalised pneumonia groups for the statistical analysis. Hospitalised pneumonia participants re-admitted to hospital within 3-months post-recruitment will be excluded from the control group in the case-control analysis.

Continuous variables will be summarised by means and SD, or medians and IQR if the distribution exhibits skew. Beta diversity of the NPR will be quantified using Bray-Curtis dissimilarities and compared across study groups at single study time points using PERMANOVA, with permutations restricted within participants for comparisons between time points. Alpha diversity (measured using the Shannon Diversity Index), relative abundance of individual ARGs and ARG classes in the NPR, and relative abundance of bacterial species in the NPM, will be compared across study groups using the Kruskal-Wallis test with Dunn’s post-hoc pairwise comparisons, and between time-points within participants using the Wilcoxon signed-rank test. Differences in incidence of adverse health outcomes will be compared across study groups using the Wilcoxon Rank Sum test. False discovery rate correction will be used to adjust for multiple comparisons.

Primary endpoint analyses will only include participants with complete follow-up data. Analyses for secondary and exploratory outcomes will include all participants. Missing data will be fully described.

## ETHICS AND DISEMINATION

### Ethical approvals

The study has been approved by the Malawi College of Medicine Research Ethics Committee (COMREC; Ref: P.10/24-1200) and Liverpool School of Tropical Medicine (LSTM; Ref: 24-076).

### Informed consent process

The study will only recruit children whose parents/legally acceptable representatives have the capacity to provide written informed consent. In situations where the parent/legally acceptable representative is illiterate, an impartial witness will be asked to verify the participant’s understanding and will also sign on the ICF; the participant’s parent/legally acceptable representative will acknowledge their agreement with a thumbprint. Parents/legally acceptable representatives will receive both verbal and written information about the study, and be given the opportunity to ask questions, reflect on the information provided and come to a decision, before accepting to take part. Parents/legally acceptable representatives of participants will also be informed of their right to withdraw consent at any point without the need to provide a reason, and without penalty.

### Dissemination policy and plans

The study results will be shared with local stakeholders, local and international research ethics committees, and the Malawi Ministry of Health after study completion and data analysis. Interim analyses may be presented at relevant local and international scientific meetings. All research outputs from this study will be published on open-source platforms or in open-source peer-reviewed journals. The study findings will be shared with participating communities, to ensure that research outputs benefit the local population. Open access publication and sharing of curated metagenomic data on the GitHub repository will ensure long-term data preservation.

## Data Availability

This is a study protocol, so does not include any data.

## Acknowledgements

The authors thank the MLW laboratory management team, led by Brigitte Denis and George Selemani, and Jeromy Kawaye and the MLW data management team.

## Authors contributions

Contributors LOC, RH and NF conceived the study. LOC, RH, NF, BKA and CC designed the study with contributions from CM, AK and MH. LOC wrote the first draft. All authors have commented on the manuscript. All authors have read and approved the final manuscript.

## REFERENCES

1. O’Neill J. Tackling drug-resistant infections globally: Final report and recommendations. Arch Pharm Pract. 2016;7(3):110. doi:10.4103/2045-080x.186181

2. Murray CJ, Ikuta KS, Sharara F, Swetschinski L, Robles Aguilar G, Gray A, et al. Global burden of bacterial antimicrobial resistance in 2019: a systematic analysis. The Lancet. 2022 Feb 12;399(10325):629–55. doi:10.1016/S0140-6736(21)02724-0 PubMed PMID: 35065702.

3. World Health Organisation. World Health Organisation [Internet]. World Health Organisation; 2022 [cited 2026 Apr 18]. Child mortality (under 5 years). Available from: https://www.who.int/news-room/fact-sheets/detail/child-mortality-under-5-years

4. Siegel SJ, Weiser JN. Mechanisms of Bacterial Colonization of the Respiratory Tract. Annu Rev Microbiol. 2015;69:425–44. doi:10.1146/annurev-micro-091014-104209 PubMed PMID: 26488280; PubMed Central PMCID: PMC4760621.

5. Man WH, van Houten MA, Mérelle ME, Vlieger AM, Chu MLJN, Jansen NJG, et al. Bacterial and viral respiratory tract microbiota and host characteristics in children with lower respiratory tract infections: a matched case-control study. Lancet Respir Med. 2019 May 1;7(5):417–26. doi:10.1016/S2213-2600(18)30449-1

6. Schmieder R, Edwards R. Insights into antibiotic resistance through metagenomic approaches. Future Microbiol. 2012 Jan;7(1):73–89. doi:10.2217/fmb.11.135 PubMed PMID: 22191448.

7. Wright GD. The antibiotic resistome. Expert Opin Drug Discov. 2010;5(8):779–88. doi:10.1517/17460441.2010.497535 PubMed PMID: 22827799.

8. Tantivitayakul P, Lapirattanakul J, Vichayanrat T, Muadchiengka T. Antibiotic Resistance Patterns and Related Mobile Genetic Elements of Pneumococci and β-Hemolytic Streptococci in Thai Healthy Children. Indian J Microbiol. 2016 Dec 1;56(4):417–25. doi:10.1007/s12088-016-0607-3

9. Kalizang’oma A, Chaguza C, Gori A, Davison C, Beleza S, Antonio M, et al. Streptococcus pneumoniae serotypes that frequently colonise the human nasopharynx are common recipients of penicillin-binding protein gene fragments from streptococcus mitis. Microb Genomics. 2021;7(9). doi:10.1099/mgen.0.000622 PubMed PMID: 34550067.

10. O’Connor L, Heyderman R. The challenges of defining the human nasopharyngeal resistome. Trends Microbiol. 2023 Mar. doi:10.1016/j.tim.2023.02.008 PubMed PMID: 36967247.

11. Makori TO, Gicheru ET, Mburu MW, Sada MS, Nyawa O, Mutunga M, et al. Nasopharyngeal Microbiome Composition and its Clinical Correlates in Children Hospitalized with Severe Pneumonia in East Africa. J Infect Dis. 2026 Sep 15;234(3):475–83. doi:10.1093/infdis/jiag093

12. Tizifa TA, Kabaghe AN, McCann RS, Nkhono W, Mtengula S, Takken W, et al. Incidence of clinical malaria, acute respiratory illness, and diarrhoea in children in southern Malawi: a prospective cohort study. Malar J. 2021;20(1):473. doi:10.1186/s12936-021-04013-5

13. World Health Organisation. Cough or Difficulty in Breathing. In: Pocket book of hospital care for children: guidelines for the management of common childhood illnesses 2nd edition [Internet]. 2nd edn. Geneva, Switzerland: World Health Organisation; 2013 [cited 2026 Apr 22]. p. 75–103. Available from: https://iris.who.int/server/api/core/bitstreams/8f110da0-22e6-4ef1-90e4-c9f1b7daa363/content

14. Gallagher J, Chisale M, Das S, Drew RJ, Glezeva N, Wildes DM, et al. Aetiology and severity of childhood pneumonia in primary care in Malawi: a cohort study. BMJ Open. 2021 Jul 1;11(7):e046633. doi:10.1136/bmjopen-2020-046633 PubMed PMID: 34326047.

15. Ginsburg AS, Mvalo T, Nkwopara E, McCollum ED, Ndamala CB, Schmicker R, et al. Placebo vs Amoxicillin for Nonsevere Fast-Breathing Pneumonia in Malawian Children Aged 2 to 59 Months: A Double-blind, Randomized Clinical Noninferiority Trial. JAMA Pediatr. 2019 Jan 1;173(1):21–8. doi:10.1001/jamapediatrics.2018.3407 PubMed PMID: 30419120; PubMed Central PMCID: PMC6583426.

16. Brinkhoff T. City Population [Internet]. [cited 2026 Apr 21]. Blantyre City (Malawi) - Population Statistics, Charts, Map and Location. Available from: https://www.citypopulation.de/en/malawi/sub/admin/MW315_blantyre_city/

17. Brinkhoff T. City Population [Internet]. National Statistical Office of Malawi; [cited 2026 Apr 21]. City Population Malawi: Administrative Division (Regions and Districts) - Population Statistics, Charts and Map. Available from: https://www.citypopulation.de/en/malawi/admin/

18. Swarthout TD, Fronterre C, Lourenço J, Obolski U, Gori A, Bar-Zeev N, et al. High residual carriage of vaccine-serotype Streptococcus pneumoniae after introduction of pneumococcal conjugate vaccine in Malawi. Nat Commun. 2020;11(1):1–12. doi:10.1038/s41467-020-15786-9 PubMed PMID: 32376860.

19. Flynn M, Dooley J. The microbiome of the nasopharynx. J Med Microbiol. 2021;70(6):001368. doi:10.1099/jmm.0.001368 PubMed PMID: 34165422.

20. O’Brien KL, Baggett HC, Brooks WA, Feikin DR, Hammitt LL, Higdon MM, et al. Causes of severe pneumonia requiring hospital admission in children without HIV infection from Africa and Asia: the PERCH multi-country case-control study. The Lancet. 2019 Aug 31;394(10200):757–79. doi:10.1016/S0140-6736(19)30721-4

21. Skovbjerg S, Brown C, Mataya AA, Msefula J, Bonomali F, Banda F, et al. Urine bioassay optimisation to assess the association between antimicrobial exposure, pneumococcal carriage and antimicrobial resistance among hospitalised children in Malawi. BMC Infect Dis. 2025 Oct 13;25(1):1291. doi:10.1186/s12879-025-11871-w PubMed PMID: 41083965; PubMed Central PMCID: PMC12516879.

22. Ngari MM, Obiero C, Mwangome MK, Nyaguara A, Mturi N, Murunga S, et al. Mortality during and following hospital admission among school-aged children: a cohort study. Wellcome Open Res. 2020;5:234. doi:10.12688/wellcomeopenres.16323.2 PubMed PMID: 33195820.

23. Akinbami FO, Orimadegun AE, Tongo OO, Okafor OO, Akinyinka OO. Detection of fever in children emergency care: Comparisons of tactile and rectal temperatures in Nigerian children. BMC Res Notes. 2010;3. doi:10.1186/1756-0500-3-108

24. Odinaka K, Edelu B, Nwolisa E, Amamilo I, Okolo S. Accuracy of subjective assessment of fever by Nigerian mothers in under-5 children. Niger Med J. 2014;55(4):338. doi:10.4103/0300-1652.137226

25. Connon R, George EC, Olupot-Olupot P, Kiguli S, Chagaluka G, Alaroker F, et al. Incidence and predictors of hospital readmission in children presenting with severe anaemia in Uganda and Malawi: a secondary analysis of TRACT trial data. BMC Public Health. 2021;21(1):1480. doi:10.1186/s12889-021-11481-6

26. Peterson I, Bar-Zeev N, Kennedy N, Ho A, Newberry L, SanJoaquin MA, et al. Respiratory virus-associated severe acute respiratory illness and viral clustering in Malawian children in a setting with a high prevalence of HIV infection, malaria, and malnutrition. J Infect Dis. 2016 Dec 1;214(11):1700–11. doi:10.1093/infdis/jiw426 PubMed PMID: 27630199.

27. Cao Q, Sun X, Rajesh K, Chalasani N, Gelow K, Katz B, et al. Effects of Rare Microbiome Taxa Filtering on Statistical Analysis. Front Microbiol. 2021 Jan 12;11:607325. doi:10.3389/fmicb.2020.607325 PubMed PMID: 33510727; PubMed Central PMCID: PMC7835481.

28. Zhou H, Larkin PMK, Zhao D, Ma Q, Yao Y, Wu X, et al. Clinical Impact of Metagenomic Next-Generation Sequencing of Bronchoalveolar Lavage in the Diagnosis and Management of Pneumonia: A Multicenter Prospective Observational Study. J Mol Diagn. 2021 Oct 1;23(10):1259–68. doi:10.1016/j.jmoldx.2021.06.007

29. Zhang Z, Zhang Q, Wang T, Xu N, Lu T, Hong W, et al. Assessment of global health risk of antibiotic resistance genes. Nat Commun. 2022 Dec 1;13(1). doi:10.1038/s41467-022-29283-8 PubMed PMID: 35322038.

30. Hou J, Song Y, Leung ASY, Tang MF, Shi M, Wang EY, et al. Temporal Dynamics of the Nasopharyngeal Microbiome and its Relationship with Childhood Asthma Exacerbation. Microbiol Spectr. 2022 Jun;10(3):e0012922. doi:10.1128/spectrum.00129-22 PubMed PMID: 35546575.

31. ODK - Collect data anywhere [Internet]. Get ODK Inc.; [cited 2026 Apr 23]. Available from: https://getodk.org

32. Sonego M, Pellegrin MC, Becker G, Lazzerini M. Risk factors for mortality from acute lower respiratory infections (ALRI) in children under five years of age in low and middle-income countries: a systematic review and meta-analysis of observational studies. PloS One. 2015;10(1):e0116380. doi:10.1371/journal.pone.0116380 PubMed PMID: 25635911; PubMed Central PMCID: PMC4312071.

33. Teo SM, Mok D, Pham K, Kusel M, Serralha M, Troy N, et al. The infant nasopharyngeal microbiome impacts severity of lower respiratory infection and risk of asthma development. Cell Host Microbe. 2015;17(5):704–15. doi:10.1016/j.chom.2015.03.008 PubMed PMID: 25865368.

34. Biesbroek G, Tsivtsivadze E, Sanders EAM, Montijn R, Veenhoven RH, Keijser BJF, et al. Early respiratory microbiota composition determines bacterial succession patterns and respiratory health in children. Am J Respir Crit Care Med. 2014 Dec 1;190(11):1283–92. doi:10.1164/rccm.201407-1240OC PubMed PMID: 25329446.

35. Bosch AATM, Levin E, van Houten MA, Hasrat R, Kalkman G, Biesbroek G, et al. Development of Upper Respiratory Tract Microbiota in Infancy is Affected by Mode of Delivery. EBioMedicine. 2016 Jul;9:336–45. doi:10.1016/j.ebiom.2016.05.031 PubMed PMID: 27333043.

36. King C, Banda M, Bar-Zeev N, Beard J, French N, Makwenda C, et al. Care-seeking patterns amongst suspected paediatric pneumonia deaths in rural Malawi. Gates Open Res. 2020;4:178. doi:10.12688/gatesopenres.13208.2 PubMed PMID: 33537557.

37. Harris M, Clark J, Coote N, Fletcher P, Harnden A, McKean M, et al. British Thoracic Society guidelines for the management of community acquired pneumonia in children: update 2011. Thorax. 2011 Oct;66 Suppl 2:ii1–23. doi:10.1136/thoraxjnl-2011-200598 PubMed PMID: 21903691.

38. Ferdous T, Jiang L, Dinu I, Groizeleau J, Kozyrskyj AL, Greenwood CMT, et al. The rise to power of the microbiome: power and sample size calculation for microbiome studies. Mucosal Immunol. 2022 Jun;15(6):1060–70. doi:10.1038/s41385-022-00548-1 PubMed PMID: 35869146.

39. Willmann M, Vehreschild MJGT, Biehl LM, Vogel W, Dörfel D, Hamprecht A, et al. Distinct impact of antibiotics on the gut microbiome and resistome: A longitudinal multicenter cohort study. BMC Biol. 2019;17(1):1–18. doi:10.1186/s12915-019-0692-y PubMed PMID: 31533707.

40. Manenzhe RI, Dube FS, Wright M, Lennard K, Mounaud S, Lo SW, et al. Characterization of Pneumococcal Colonization Dynamics and Antimicrobial Resistance Using Shotgun Metagenomic Sequencing in Intensively Sampled South African Infants. Front Public Health. 2020;8:543898. doi:10.3389/fpubh.2020.543898 PubMed PMID: 33072693.

41. Manenzhe RI, Dube FS, Wright M, Lennard K, Zar HJ, Mounaud S, et al. Longitudinal changes in the nasopharyngeal resistome of South African infants using shotgun metagenomic sequencing. PloS One. 2020 Apr 22;15(4):e0231887–e0231887. doi:10.1371/journal.pone.0231887

42. Allemann A, Kraemer JG, Korten I, Ramsey K, Casaulta C, Wüthrich D, et al. Nasal Resistome Development in Infants With Cystic Fibrosis in the First Year of Life. Front Microbiol. 2019;10:212.

43. Cason C, D’Accolti M, Campisciano G, Soffritti I, Ponis G, Mazzacane S, et al. Microbial Contamination in Hospital Environment Has the Potential to Colonize Preterm Newborns’ Nasal Cavities. Pathog Basel Switz. 2021 May 17;10(5):615. doi:10.3390/pathogens10050615

44. Keenan JD, Chin SA, Amza A, Kadri B, Nassirou B, Cevallos V, et al. The Effect of Antibiotic Selection Pressure on the Nasopharyngeal Macrolide Resistome: A Cluster-randomized Trial. Clin Infect Dis Off Publ Infect Dis Soc Am. 2018 Nov;67(11):1736–42. doi:10.1093/cid/ciy339 PubMed PMID: 29897440.

45. Bojang A, Baines SL, Camara B, Guerillot R, Donovan L, Marqués RS, et al. Impact of Intrapartum Oral Azithromycin on the Acquired Macrolide Resistome of Infants’ Nasopharynx: A Randomized Controlled Trial. Clin Infect Dis. 2020 Dec 15;71(12):3222–5. doi:10.1093/cid/ciaa609

46. García-Rodríguez JA, Fresnadillo Martínez MJ. Dynamics of nasopharyngeal colonization by potential respiratory pathogens. J Antimicrob Chemother. 2002 Dec;50 Suppl S:59–73. doi:10.1093/jac/dkf506 PubMed PMID: 12556435.

47. Doan T, Yan D, Arzika AM, Abdou A, Maliki R, Aichatou B, et al. Mass azithromycin distribution and antibiotic resistance in the gut and nasopharynx: a cluster-randomized trial. Nat Med. 2026 Mar;32(3):859–68. doi:10.1038/s41591-026-04217-9

48. FigureLabs [Internet]. [cited 2026 Oct 29]. Available from: www.figurelabs.ai

49. Gemini [Internet]. Google; [cited 2026 Oct 29]. Available from: https://gemini.google.com/

